# Feasibility of Deep Learning Segmentation in Anterior Quadratus Lumborum Ultrasound Imaging: Challenges in Identifying the Injection Plane

**DOI:** 10.1101/2025.10.29.25339036

**Authors:** Daniel Bidstrup, Anuj Pareek, Jens Børglum

## Abstract

**Background:** Accurate identification of sonoanatomy is essential for successful anterior quadratus lumborum (QL) block performance. Prior deep learning work in QL ultrasound imaging has focused on segmentation of anatomically distinct muscular and osseous structures. Automated segmentation of the anterior QL block injection plane, however, has not been described.

**Methods:** In this single-center prospective imaging study, 82 simulated anterior QL block ultrasound recordings from 42 healthy volunteers yielded 460 annotated frames across six classes: vertebral body (L3/L4), posterior renal fascia, transverse abdominal muscle, quadratus lumborum muscle, psoas major muscle, and the anterior QL block injection plane. A 2D nnU-Net model was trained using five-fold cross-validation and tested on 23 unseen clinical images acquired during routine anterior QL block procedures. Performance was assessed using Dice similarity coefficients.

**Results:** The mean Dice score across all classes was 0.62. Segmentation performance was highest for the vertebral body (0.90) and psoas major muscle (0.85), moderate for the quadratus lumborum muscle (0.69) and transverse abdominal muscle (0.51), and lower for the posterior renal fascia (0.35) and injection plane (0.38).

**Conclusions:** Deep learning was feasible for segmentation of major anatomical landmarks in anterior QL ultrasound images, but performance remained limited for the interfascial injection plane. Further development and external validation are required before clinical application.

## INTRODUCTION

The ultrasound-guided quadratus lumborum (QL) block is a regional anesthesia technique that has gained increasing popularity for postoperative pain management. There are different approaches to perform a QL block, including the anterior, lateral, and posterior approach. Each block technique has its unique application guidelines and possible benefits.

Introduced in 2013, the ultrasound-guided anterior QL block was shown in a cadaveric study to enable injectate to spread from its lumbar site of injection cranially towards the diaphragmatic openings and further into the thoracic paravertebral space, and thereby reaching the thoracic sympathetic trunk and the lower thoracic nerves[1]. The anterior QL block also simultaneously anesthetizes the subcostal, ilioinguinal, and iliohypogastric nerves, suggesting its ability to provide both somatic and visceral pain relief[1]. Clinical trials have demonstrated reduced postoperative opioid consumption following cesarean section, laparoscopic nephrectomy, and percutaneous nephrolithotomy[2–4]. A recent systematic review with meta-analyses further supports its analgesic efficacy when performed correctly[5].

Despite these reported benefits, the effectiveness of the anterior QL block depends on accurate identification of the relevant sonoanatomy. The QL block injection plane lies deep within the posterior abdominal wall and is embedded in a multilayered thoracolumbar fascial complex. The intended injection site represents an interfascial plane between the quadratus lumborum and psoas major muscles, bounded by the anterior layer of the thoracolumbar fascia and the transversalis fascia. While the surrounding region is delineated by recognizable muscular and fascial structures, the injection target itself lacks intrinsic sonographic borders and is defined relationally by adjacent anatomical landmarks.

In parallel with the growing adoption of ultrasound-guided regional anesthesia, there has been increasing interest in the use of Artificial Intelligence (AI) for assistance in image interpretation. Prior studies have demonstrated AI-based needle detection and segmentation of superficial nerve block anatomy[6–8]. Recently, a deep learning model has been described for segmentation of the quadratus lumborum muscle, vertebral bones, and transverse abdominal muscle group on ultrasound images[9]. That work focused on anatomically distinct muscular and osseous structures. In contrast, automated segmentation of the anterior QL block injection plane has not been described.

In this feasibility study, we aimed to develop and evaluate a convolutional neural network for segmentation of key sonoanatomical landmarks relevant to the anterior QL block. In addition to well-defined structures such as vertebral bodies and major muscles, we specifically included the interfascial injection plane of the anterior QL block as a segmentation target. This procedural interfascial plane has not previously been addressed in AI-based segmentation models.

## METHODS

### Training Data

In this single-center, prospective imaging study, we utilized video recordings of simulated (without needle placement) ultrasound-guided anterior QL block procedures at vertebral level L3/L4. All ultrasound recordings were obtained by an experienced consultant anesthesiologist (JB) with extensive clinical and research experience in quadratus lumborum block techniques. The recordings were acquired from 42 healthy medical students who volunteered while on clinical training at our department (see Table 1). Of these, 40 volunteers were scanned bilaterally and two were scanned unilaterally, resulting in a total of 82 unique recordings. Each ultrasound recording had a 30-second duration, which was sufficient to capture dynamic anatomical changes across respiratory cycles. These dynamic changes guide the ultrasound operator for better anatomical recognition in daily clinical practice. Important anatomical structures like the para- and perinephric fat, as well as the posterior renal fascia, change position and appearance during respiratory cycles[2]. This approach allowed us to collect a broader range of sonoanatomical variation and thereby helped us obtain more useful training data from each volunteer.

**Table 1.** Descriptive characteristics of healthy volunteers (training data).

|  | Age | Height (cm) | Weight (kg) | BMI (kg/m<sup>2</sup>) | Recordings | Annotated frames |
| --- | --- | --- | --- | --- | --- | --- |
| Overall (n = 42) | 30 (9) | 175 (14) | 72 (20) | 23.7 (5.2) | 82 | 460 |
| Female (n = 25) | 29 (5) | 171 (9) | 65 (12) | 22.7 (6.0) | 48 | 280 |
| Male (n = 17) | 32 (13) | 185 (10) | 82 (13) | 24.3 (3.8) | 34 | 180 |

Values are median (interquartile range) unless otherwise indicated. BMI, body mass index.

### Training Data Selection

Each 30-second ultrasound video consisted of 360 image frames. From each video, 5–15 representative frames were carefully selected, yielding a total dataset of 460 images with a resolution of 784 × 580 pixels. Frame selection was based on two predefined criteria: (1) clear visualization of essential sonoanatomical landmarks relevant to the anterior QL block, and (2) variability in anatomical appearance to capture differences encountered in clinical practice. This approach ensured a broad representation of the anterior QL block sonoanatomy.

### Annotation and Labeling

A fifth-year resident in anesthesiology (DB) annotated all images, providing detailed ground truth labels for six different classes. These labels included the following sonoanatomic classes: 1) the vertebral body of L3 or L4 and their respective transverse process, 2) posterior renal fascia, 3) transverse abdominal muscle, 4) quadratus lumborum muscle, 5) psoas major muscle, and 6) the injection plane for the anterior QL block in the fascial interspace between the QL and psoas major muscle posterior to the transversalis fascia. In cases where uncertainty arose during annotation, the corresponding images and labels were reviewed by JB, whose assessment served as the final determination. JB did not perform independent re-annotation of the entire dataset but served as a senior reviewer for cases requiring clarification. The annotation and labelling were done using the Labelbox online tool[10].

### AI Model Development

We set up our AI model as the nnU-Net[11] self-configuring method for semantic segmentation of biomedical images in PyTorch[12]. The nnU-Net pipeline makes use of the 2D U-net architecture[13] which has consistently demonstrated robust performance on segmentation tasks. The nnU-Net pipeline automatically normalizes the input images and performs data augmentation including rotations, scaling, and Gaussian noise.. The nnU-Net was trained on an NVIDIA L4 24GB GPU using five-fold cross-validation meaning the data was split into an 80% training set (368 images) and 20% validation set (92 images) for each fold. Each fold was trained for 1000 epochs, with 250 minibatches per epoch and a minibatch size of six. The model was optimized using stochastic gradient descent with Nesterov momentum and a decaying learning rate[14]. The loss function combined the sum of Cross-Entropy loss and Dice loss to balance pixel-wise classification with overlap-based segmentation performance. Model performance was evaluated using the average Dice score across all six label classes, and following five-fold cross-validation, the final model was selected as the best-performing ensemble from the trained folds.

### Performance Evaluation

The effectiveness of our AI model was quantitatively assessed using the Dice score metric[15]. The Dice score, also known as the Dice Similarity Coefficient, is a statistical measure used to gauge the similarity between two sets[16, 17]. In our study, it was used to evaluate the agreement between the AI-predicted segmentations and the manually annotated sonoanatomy in the ultrasound images. We categorized Dice similarity coefficients into five bins; low [0.00, 0.20), low-moderate [0.20, 0.40), moderate [0.40, 0.60), moderate-high [0.60, 0.80), and high [0.80, 1.00] consistent with prior work in biomedical image segmentation[18].

### Test Data and Evaluation

To evaluate model performance in a clinical context, we used a test set consisting of single ultrasound images from 23 patients who received an anterior QL block as part of routine perioperative care. Twenty-one patients underwent block placement following elective cesarean section, and two patients received the block prior to laparoscopic nephrectomy (Table 2). All test-set images were acquired by JB during routine clinical practice. For each patient, a single ultrasound image was obtained at the time of block performance, representing the sonographic view used for anatomical assessment prior to needle advancement. Image acquisition reflected standard clinical workflow, in which block placement is performed under the best achievable visualization of relevant anatomical landmarks.

**Table 2.** Descriptive characteristics of patients (test data).

|  | Age | Height (cm) | Weight (kg) | BMI (kg/m<sup>2</sup>) | Annotated frames |
| --- | --- | --- | --- | --- | --- |
| Overall (n = 23) | 33 (8) | 168 (10) | 77 (18) | 32.5 (9.5) | 23 |
| Female (n = 23) | 33 (8) | 168 (10) | 77 (18) | 32.5 (9.5) | 23 |
| Male (n = 0) | — | — | — | — | 0 |

Values are median (interquartile range) unless otherwise indicated. BMI, body mass index.

All 23 test images were labeled and annotated by DB and subsequently reviewed by JB to provide ground truth labels. The final trained nnU-Net model ensemble was used to predict segmentation labels for each of the 23 images.

The model performance on these unseen images was then quantitatively assessed for each sonoanatomic class using the Dice score by comparing the predicted labels with our ground truth labels.

## RESULTS

We achieved an average Dice score of 0.62 across all anatomical classes in the test set (see Table 3). This value reflects the overall accuracy of the model in identifying and segmenting the relevant sonoanatomical landmarks for the anterior QL block. Dice scores for the individual label classes are presented in Table 3.

**Table 3.** Performance of the deep learning model on the clinical test set (n=23).

| Segmentation target | Dice score |
| --- | --- |
| Mean across all classes | 0.62 |
| *Individual segmentation classes* |  |
| Vertebral body of L3 or L4 | 0.90 |
| Psoas major muscle | 0.85 |
| Quadratus lumborum muscle | 0.69 |
| Transverse abdominal muscle | 0.51 |
| Posterior renal fascia | 0.35 |
| Anterior QL block injection plane | 0.38 |

Dice score quantifies the spatial overlap between model-predicted and reference segmentations (range 0 [no overlap] to 1 [perfect overlap]). QL, quadratus lumborum.

Figure 1 illustrates our AI model’s segmentation performance in five selected patients by comparing ground truth labels with model predicted labels. The segmentation performance in all 23 test patients is showcased in supplemental material Figure S1A-E. The supplementary Table S1 provides a bin-wise breakdown of Dice similarity coefficients (low to high) for each segmented structure across the 23 test cases. In Patient 1, the predicted labels closely matched the reference annotations across all label classes. Patient 2 exemplifies cases in which the model showed reduced consistency in identifying the posterior renal fascia and the transverse abdominal muscle. Patient 3 shows that the anterior QL block injection plane was generally detected, although the predicted region was smaller than the reference annotation. Patients 4 and 5 demonstrate that despite achieving a Dice score of 0.69 for the quadratus lumborum muscle, the predicted label is spatially displaced in some images. Patient 5 represented an example of anatomy outside the distribution of the training data; this patient had renal cancer, which altered the local anatomy and was associated with reduced segmentation accuracy.

**Figure 1:**
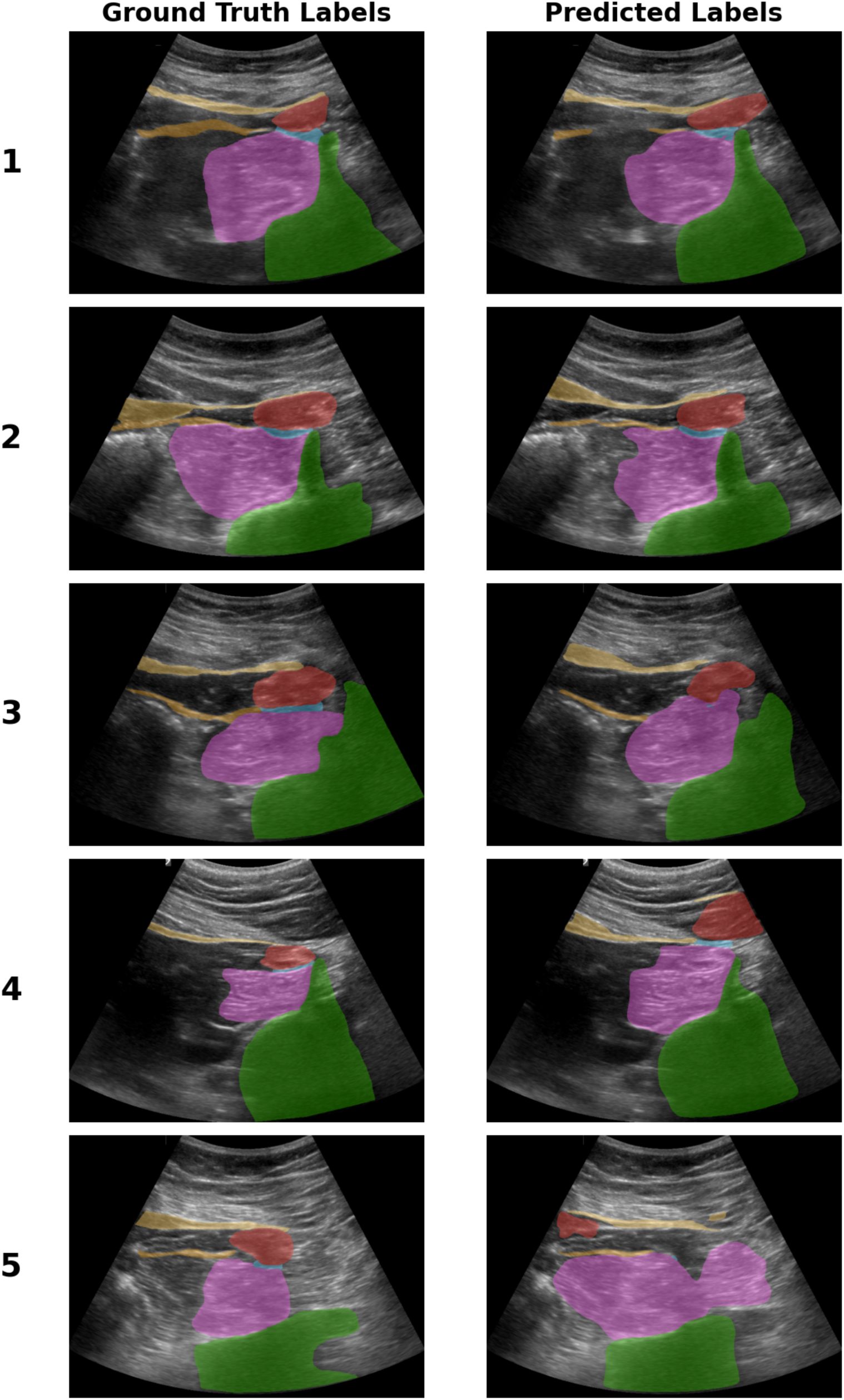
Comparison of Ground Truth and Predicted Labels for AI Model Performance on Five Selected Patients. Patient 1 shows near-perfect agreement. Patient 2 highlights challenges in identifying the posterior renal fascia and transverse abdominal muscle. Patient 3 demonstrates underestimation of the anterior QL block injection plane. Patients 4 and 5 show occasional misplacement of the quadratus lumborum muscle label despite a Dice score of 0.69. Patient 5 further illustrates poor performance on out-of-distribution sonoanatomy due to renal cancer-induced anatomical distortion. Green = Vertebral body of L3 or L4, Red = Psoas major muscle, Yellow = Transverse abdominal muscle, Brown = Posterior renal fascia, Blue = Anterior QL block injection plane.

## DISCUSSION

Recent work has explored deep learning–based segmentation in QL block ultrasound imaging, most notably a VGG16– U-Net–based model (Q-VUM) trained on 3,162 images from 112 patients[9]. This study reported high Dice performance for segmentation of structurally distinct tissues, including the quadratus lumborum muscle and vertebral elements. However, several methodological differences limit direct comparison with our study. First, Wang et al. focused exclusively on anatomically well-defined muscular and osseous structures with intrinsic sonographic borders. In contrast, we additionally attempted segmentation of the anterior QL injection plane, which represents a procedural target between the quadratus lumborum and psoas major muscles. Unlike muscle or bone, this target lacks fixed morphological boundaries and is only defined relationally by surrounding landmarks. This makes the anterior QL block injection plane a challenging area to accurately delineate. Second, the previous study was based on a substantially larger dataset with multi-expert annotation and exclusion of lower-quality images, whereas our training dataset was smaller and derived from a limited volunteer cohort. Third, segmentation of small or thin anatomical targets is known to penalize Dice-based metrics disproportionately, particularly when label boundaries are inherently ambiguous[19]. These factors should be considered when interpreting differences in reported segmentation performance.

In our test set, segmentation performance varied markedly between structures. High Dice scores were observed for anatomically distinct and morphologically stable landmarks, including the vertebral body (0.90) and the psoas major muscle (0.85). The quadratus lumborum muscle achieved a Dice score of 0.69, indicating moderate performance. Lower Dice scores were observed for thinner or less well-circumscribed structures, including the transverse abdominal muscle (0.51), the posterior renal fascia (0.35), and the anterior QL block injection plane (0.38). The comparatively low Dice score for the injection plane reflects the inherent difficulty of segmenting a relational procedural target without intrinsic sonographic borders. Similarly, the posterior renal fascia is a thin and variably visualized structure, which likely contributed to reduced overlap metrics[19].

Differences between the training and test populations may also have influenced performance. The volunteers in the training dataset had a median BMI of 23.7 (IQR 5.2), whereas the test cohort had a median BMI of 32.5 (IQR 9.5), largely due to a high proportion of pregnant patients. Increased adiposity and pregnancy-related anatomical changes may alter tissue contrast and landmark visualization on ultrasound, introducing a domain shift between training and test data. In the two oncologic cases, local anatomical distortion further altered landmark appearance, representing anatomically challenging conditions not present in the training cohort.

Overall, the model demonstrated robust performance for large, well-defined anatomical structures and reduced performance for small or interfascial targets. These findings underscore that segmentation accuracy is structure-dependent and that fascial-plane targets represent a fundamentally more challenging task than segmentation of discrete muscular or osseous landmarks.

This study should be interpreted as a feasibility analysis rather than a clinically deployable decision-support system. The injection plane Dice score of 0.38 indicates that automated identification of this procedural target remains limited in accuracy and would not be sufficient for independent clinical guidance. However, the ability to reliably segment major anatomical landmarks suggests that deep learning may assist the operator in anatomical orientation within the anterior QL region.

The potential role of AI in ultrasound-guided regional anesthesia is evolving. AI-based tools may eventually support anatomical recognition, educational training, or real-time visualization enhancement. However, translation to clinical implementation requires larger, multi-center datasets, evaluation of interobserver variability in annotation, and validation across diverse patient populations.

In summary, segmentation performance in anterior QL ultrasound imaging differs substantially between structurally distinct landmarks and relational interfascial targets. While deep learning demonstrates feasibility for segmentation of major anatomical structures, accurate identification of the injection plane remains a significant technical challenge. These findings provide insight into the anatomical and methodological factors that influence AI-based segmentation performance in deep interfascial regional anesthesia blocks.

### Limitations

Several limitations should be considered when interpreting these findings. Our training dataset was small, derived from a limited number of healthy volunteers, and included multiple frames from the same recordings, which may limit generalizability. This concern is reinforced by the difference between the training and test populations, including higher BMI and pregnancy-related anatomical variation in the clinical cohort, which likely introduced domain shift.

Ground-truth labels were generated primarily by a single annotator, with senior review only in uncertain cases. This precluded assessment of interobserver variability and may have been particularly relevant for the anterior QL block injection plane, which lacks intrinsic sonographic borders and is therefore more vulnerable to annotation variability than discrete anatomical structures.

Our test set was also small and consisted of a single selected image per patient. This limits the precision of performance estimates and does not capture the dynamic information available during real-time ultrasound scanning.

### Future Perspectives

Future work should focus on larger and more heterogeneous datasets, multi-expert consensus annotation, and validation on full ultrasound video sequences rather than selected still images. Additional studies should also assess model robustness across different patient populations, and explore whether AI-based landmark segmentation can support anatomical orientation or training in anterior QL block procedures.

## CONCLUSION

In this feasibility study, deep learning enabled segmentation of major sonoanatomical landmarks relevant to the anterior QL block, with best performance in large and well-defined structures. Performance was substantially lower for the interfascial injection plane, underscoring the difficulty of segmenting relational procedural targets without intrinsic sonographic borders. These findings support the feasibility of AI-based landmark segmentation in anterior QL block ultrasound while highlighting the technical and methodological barriers that must be addressed before clinical application.

## Data Availability

All data produced in the present study are available from the corresponding author upon reasonable request and in compliance with Danish data protection regulations.

## DECLARATIONS

### Ethics Approval

The study was reviewed by the Regional Committee on Health Research Ethics for Region Zealand (EMN-2025-03485) who concluded that ethical approval was not required.

### Data Management

Data were processed in accordance with the EU General Data Protection Regulation (GDPR) and Danish Data Protection legislation.

### Data Availability

Training data obtained from healthy volunteers may be available from the corresponding authors upon reasonable request. Patient test data are not publicly available due to patient privacy considerations.

### Consent to Participate

All participating patients provided informed consent prior to inclusion.

### Author Contribution

Daniel Bidstrup and Anuj Pareek contributed equally to this work. Daniel Bidstrup conceived the study, designed the study protocol, annotated the training and test ultrasound datasets, contributed to data interpretation, and drafted the manuscript. Anuj Pareek designed the deep learning model architecture, implemented the data processing and training pipeline, performed model training and cross-validation, and conducted the quantitative performance evaluation. Jens Børglum provided clinical and methodological expertise in anterior quadratus lumborum block sonoanatomy, acquired all ultrasound recordings used for training and testing, contributed to study design and supervision, and served as the senior reviewer for image annotations in cases of uncertainty. All authors contributed to data interpretation, critically revised the manuscript for important intellectual content, and approved the final version to be published.

## SUPPLEMENTARY

**Table S1.** Distribution of Dice similarity coefficients across performance bins for each segmentation target.

| Segmentation target | *Dice score range* |  |  |  |  |
| --- | --- | --- | --- | --- | --- |
|  | Low | Low–Moderate | Moderate | Moderate–High | High |
|  | 0.00–0.20 | 0.20–0.40 | 0.40–0.60 | 0.60–0.80 | 0.80–1.00 |
| Vertebral body of L3 or L4 | 0 | 0 | 0 | 0 | 23 |
| Psoas major muscle | 0 | 0 | 1 | 4 | 18 |
| Quadratus lumborum muscle | 3 | 1 | 0 | 8 | 11 |
| Transverse abdominal muscle | 3 | 4 | 6 | 8 | 2 |
| Posterior renal fascia | 9 | 1 | 6 | 5 | 2 |
| Anterior QL block injection plane | 8 | 2 | 6 | 6 | 1 |

Values are the number of test cases (n = 23) falling within each Dice score range. Bin boundaries follow the convention [low, high); the highest bin is closed at 1.00. QL, quadratus lumborum.

**Figure S1.**
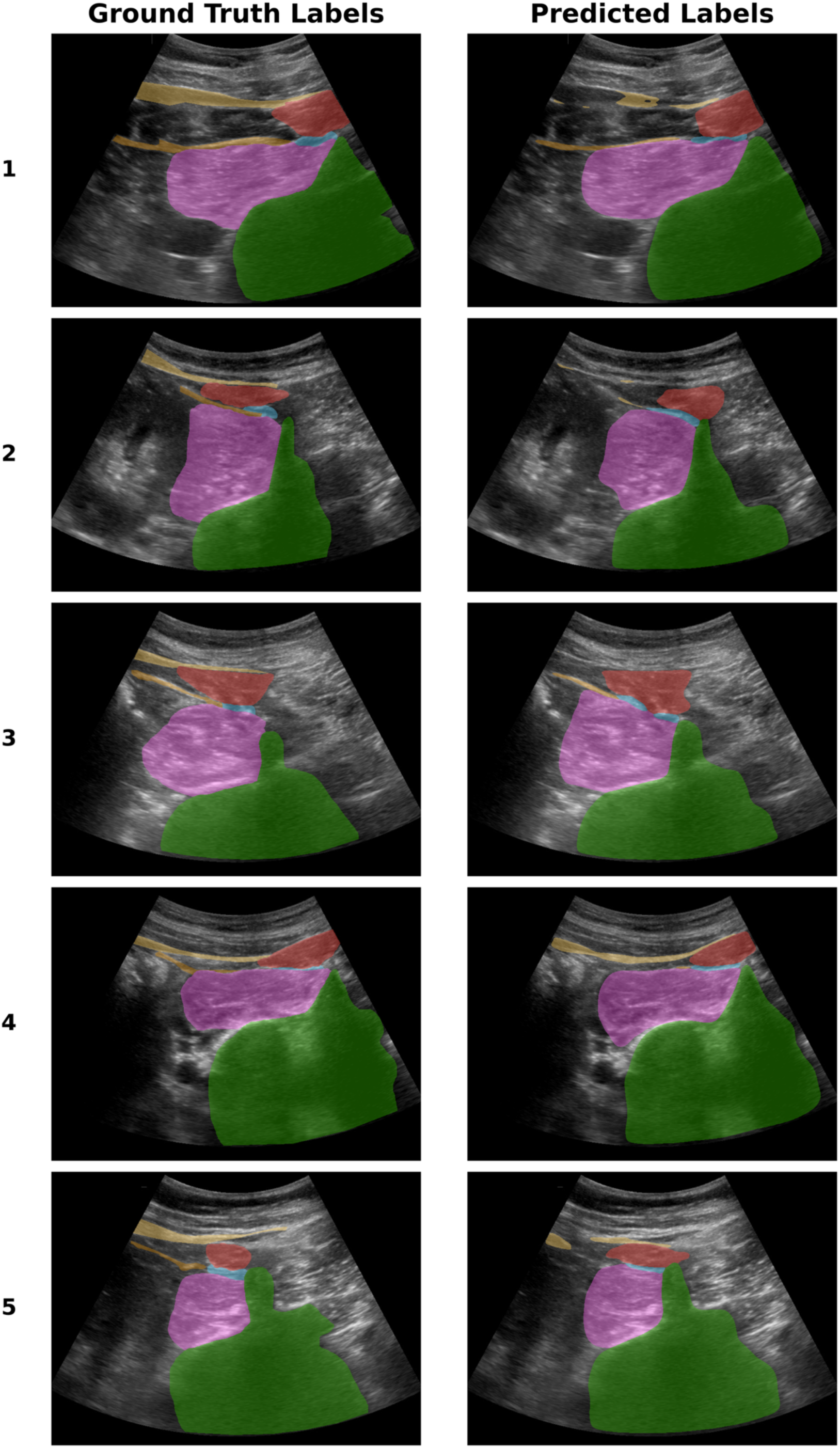
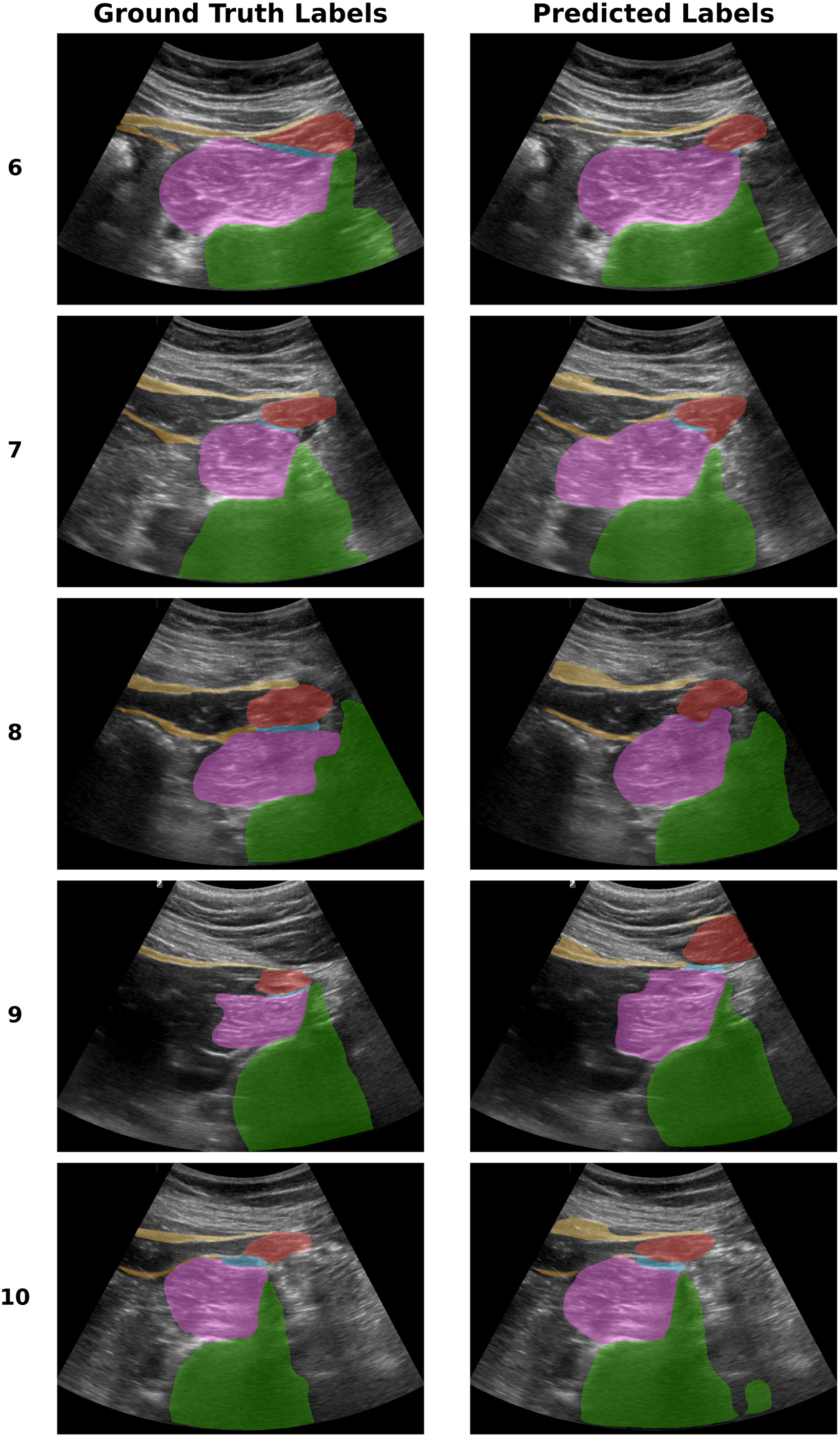
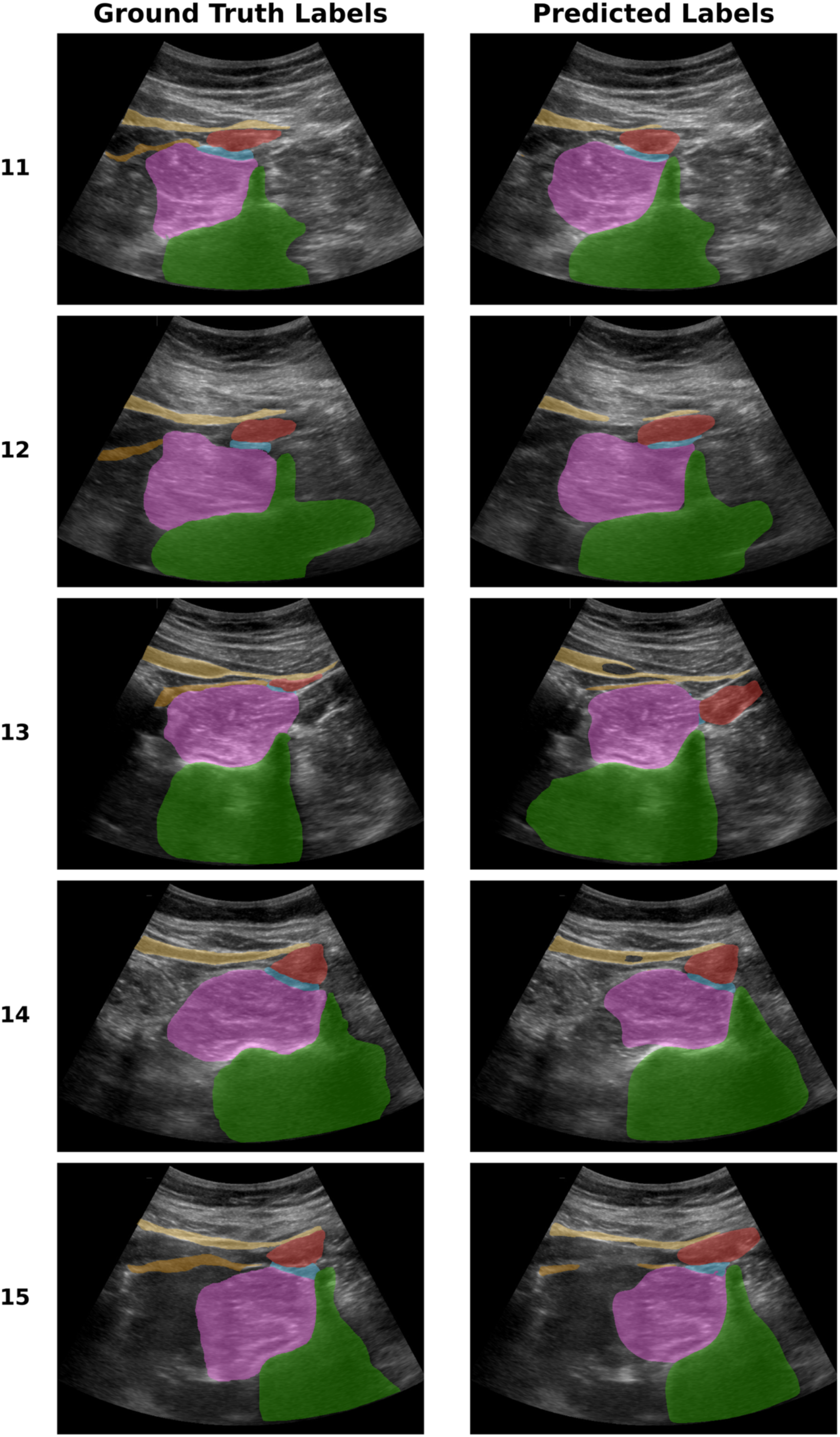
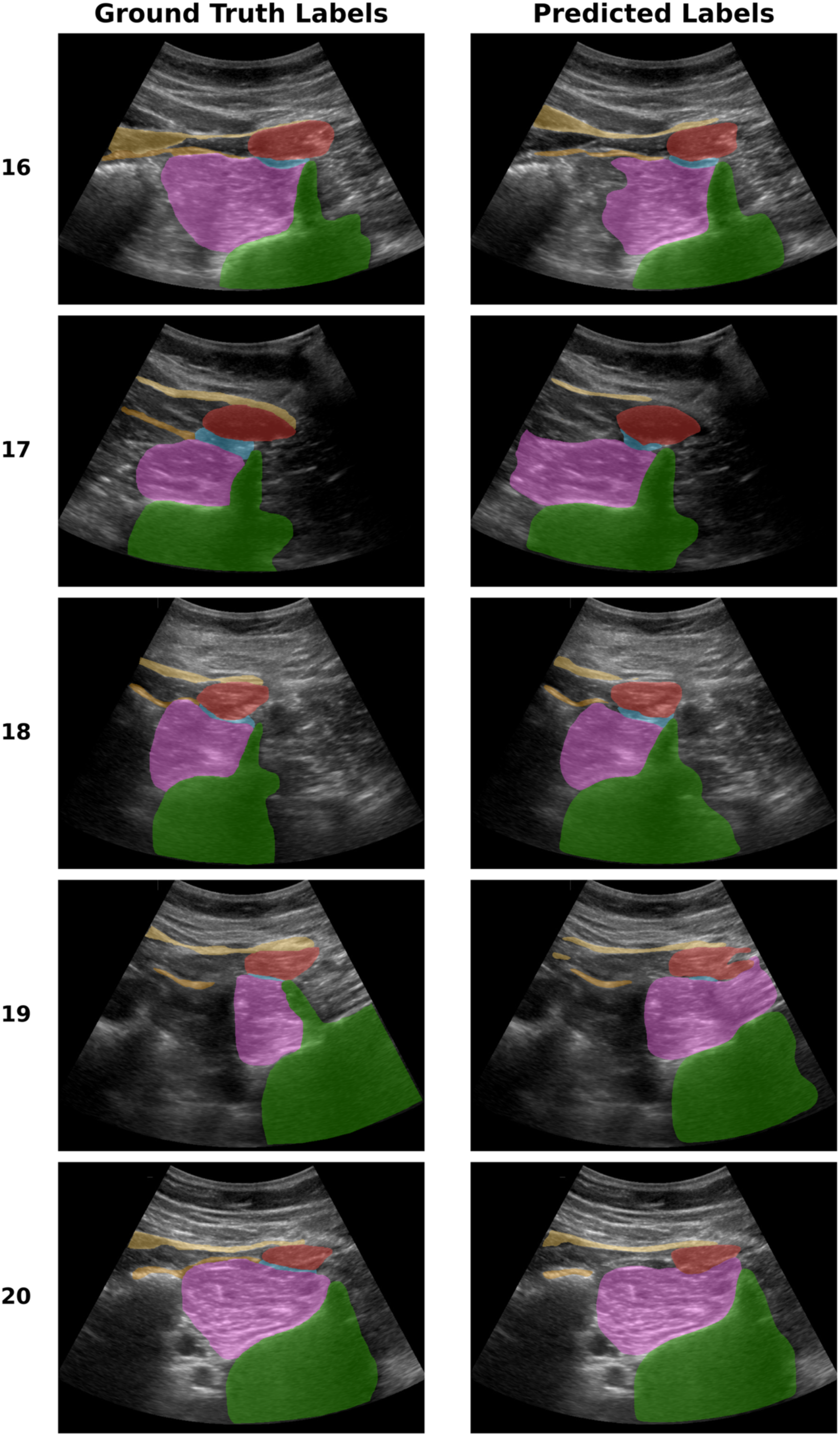
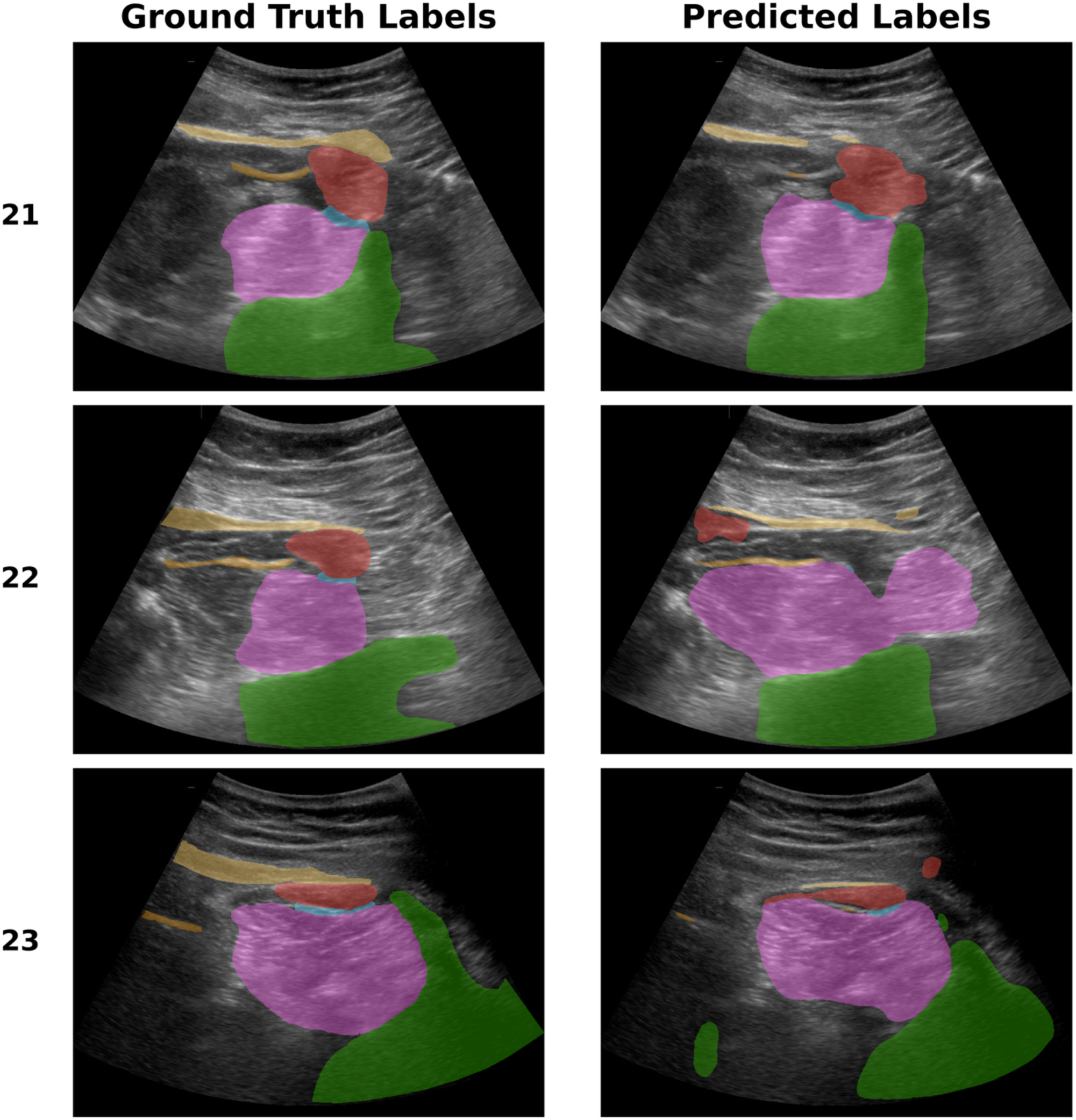
Comparison of ground-truth and AI-predicted segmentation labels in all 23 clinical test cases. Each numbered row corresponds to one patient; the left column shows the manually annotated reference labels and the right column shows the corresponding model prediction for the same ultrasound image. S1A: Patients 1–5. S1B: Patients 6–10. S1C: Patients 11–15. S1D: Patients 16–20. S1E: Patients 21–23. Color coding: Green = Vertebral body of L3 or L4, Red = Psoas major muscle, Yellow = Transverse abdominal muscle, Brown = Posterior renal fascia, Blue = Anterior QL block injection plane.

## REFERENCES

1. Dam M, Moriggl B, Hansen CK, Hoermann R, Bendtsen TF, Børglum J. The pathway of injectate spread with the transmuscular quadratus lumborum block: a cadaver study. Anesth Analg 125:303–312, 2017.

2. Dam M, Hansen C, Poulsen TD, Azawi NH, Laier GH, Wolmarans M, et al. Transmuscular quadratus lumborum block reduces opioid consumption and prolongs time to first opioid demand after laparoscopic nephrectomy. Reg Anesth Pain Med 46:18–24, 2021.

3. Dam M, Hansen CK, Poulsen TD, Azawi NH, Wolmarans M, Chan V, et al. Transmuscular quadratus lumborum block for percutaneous nephrolithotomy reduces opioid consumption and speeds ambulation and discharge from hospital: a single centre randomised controlled trial. Br J Anaesth 123:e350–e358, 2019.

4. Hansen CK, Dam M, Steingrimsdottir GE, Laier GH, Lebech M, Poulsen TD, et al. Ultrasound-guided transmuscular quadratus lumborum block for elective cesarean section significantly reduces postoperative opioid consumption and prolongs time to first opioid request: a double-blind randomized trial. Reg Anesth Pain Med 44:896–900, 2019.

5. Tanggaard K, Gronlund C, Nielsen MV, et al. Anterior quadratus lumborum blocks for postoperative pain treatment following intra-abdominal surgery: a systematic review with meta-analyses and trial sequential analyses. Acta Anaesthesiol Scand 69:e14526, 2025.

6. Viderman D, Dossov M, Seitenov S, Lee MH. Artificial intelligence in ultrasound-guided regional anesthesia: a scoping review. Front Med (Lausanne) 9:994805, 2022.

7. Mwikirize C, Nosher JL, Hacihaliloglu I. Convolution neural networks for real-time needle detection and localization in 2D ultrasound. Int J Comput Assist Radiol Surg 13:647–657, 2018.

8. Huang C, Zhou Y, Tan W, Qiu Z, Zhou H, Song Y, et al. Applying deep learning in recognizing the femoral nerve block region on ultrasound images. Ann Transl Med 7:453, 2019.

9. Wang Q, He B, Yu J, Zhang B, Yang J, Liu J, et al. Automatic segmentation of ultrasound-guided quadratus lumborum blocks based on artificial intelligence. J Imaging Inform Med 38:1362–1373, 2025.

10. Labelbox, Inc. Labelbox: Data labeling platform. San Francisco, CA. Available at: https://labelbox.com. Accessed [January 26, 2026].

11. Isensee F, Jaeger PF, Kohl SAA, Petersen J, Maier-Hein KH. nnU-Net: a self-configuring method for deep learning-based biomedical image segmentation. Nat Methods 18:203–211, 2020.

12. Paszke A, Gross S, Massa F, Lerer A, Bradbury J, Chanan G, et al. PyTorch: an imperative style, high-performance deep learning library. In: Advances in Neural Information Processing Systems 32, pp 8024-8035, 2019.

13. Ronneberger O, Fischer P, Brox T. U-net: convolutional networks for biomedical image segmentation. In: Medical Image Computing and Computer-Assisted Intervention – MICCAI 2015: 18th International Conference, Munich, Germany, October 5–9, 2015, Proceedings, Part III, Springer International Publishing, pp 234–241, 2015.

14. Sutskever I, Martens J, Dahl G, Hinton G. On the importance of initialization and momentum in deep learning. In: Dasgupta S, McAllester D, editors. Proceedings of the 30th International Conference on Machine Learning, Atlanta, GA, USA. PMLR, vol 28, pp 1139-1147, 2013.

15. Zou KH, Warfield SK, Bharatha A, Tempany CMC, Kaus MR, Haker SJ, et al. Statistical validation of image segmentation quality based on a spatial overlap index. Acad Radiol 11:178–189, 2004.

16. Dice LR. Measures of the amount of ecologic association between species. Ecology 26:297–302, 1945.

17. Sørensen T. A method of establishing groups of equal amplitude in plant sociology based on similarity of species and its application to analyses of the vegetation on Danish commons. Kongelige Danske Videnskabernes Selskab - Biologiske Skrifter 5:1–34, 1948.

18. Wilson SM, Bautista A, Yen M, Lauderdale S, Eriksson DK. Validity and reliability of four language mapping paradigms. Neuroimage Clin 16:399–408, 2017.

19. Reinke A, Tizabi MD, Baumgartner M, Eisenmann M, Heckmann-Nötzel D, Kavur AE, et al. Understanding metric-related pitfalls in image analysis validation. Nat Methods 21:182–194, 2024.

